# Experiences of Women Who Gave Birth in US Hospitals During the COVID-19 Pandemic

**DOI:** 10.1101/2020.10.15.20213504

**Authors:** Elizabeth Mollard, Amaya Wittmaack

## Abstract

The purpose of this study was to describe the experiences of women who gave birth in a US hospital during the COVID-19 pandemic. Women who gave birth between March and July 2020 completed a survey on the experience of giving birth during a pandemic. 885 women were consented and participated in the study. 22.5% of women reported hypertension, 33.8% reported anxiety, 18.6% reported depression, and 1.13% reported testing positive for COVID-19. 61% of women reported inadequate support for childbirth, and 20.5% reported that they did not feel safe giving birth in the hospital. Women who tested positive for COVID-19 were more likely to be of Asian race, have a cesarean section, not have a birth partner present, and discontinue breastfeeding before 6 weeks.

Pandemic related changes to maternity care practices may have impacted birthing women’s perceptions of safety and support in the hospital environment and affected symptoms of stress. Healthcare policy and maternity care practices should promote feelings of safety and control and overall experience for women giving birth in the hospital during a pandemic.

## Introduction

Pregnant women and maternity care practices have been significantly affected by the coronavirus-19 (COVID-19) pandemic. While the severity of COVID-19 infection in pregnant women is still being determined, pregnant women are considered a high-risk population [1-2] Early in the pandemic it was unknown whether mothers infected with COVID-19 should be separated from their infants or if they should breastfeed [3]. In the United States (US), the Center for Disease Control (CDC) recommended that infants be temporarily separated from mothers with confirmed or suspected COVID-19 infection and fed with expressed breast milk [4]. Alternatively, the World Health Organization (WHO) recommended that infected women be allowed to stay with their infants, rooming in, practicing skin-to-skin contact, and breastfeeding [5-6]. Conflicting recommendations led hospitals to create a variety of maternity care policies with varying levels of restrictions on birthing women.

Non-infected pregnant women were also greatly affected by changes to healthcare and hospital maternity care policies. Women had to face the actual and perceived threats of contracting COVID-19, while also accepting that their pregnancy and childbirth experiences would be altered. Prenatal visits moved to telehealth [7]. Universal masking procedures were enacted as well as, in some areas, universal nasopharyngeal testing for pregnant women [8] Women were uncertain if they would be able to have a birth partner, doula, or preferred provider attend their birth. When arriving at labor and delivery, even minor symptoms could be deemed a presumed positive enacting several policies to prevent infection transmission [9]. Typical best maternity care practices such as immediate skin-to-skin contact after delivery, assistance with breastfeeding in the first hour of life, and rooming-in with the infant may have been discontinued or modified [10]. Hospital stays were shortened in many areas [9]. The threat of infection transmission and changes to hospital policies made women uneasy about delivering in a hospital, provoking the professional organizations of obstetric care providers across disciplines to release a joint statement affirming that the hospital is a safe place to give birth, even during a pandemic [11].

Women who were pregnant or giving birth during the early phases of the COVID-19 pandemic faced considerable uncertainty compounded with other common potential physical and mental health comorbidities that can occur in pregnancy. Pre-pandemic research has shown that pregnant women are vulnerable to elevated levels of anxiety, depression, and other health problems such as hypertension or diabetes [12-16]. Little is known about how being pregnant or delivering a baby during a pandemic, even when not infected with COVID-19, may have affected expecting and new mothers in the US. The purpose of this study was to describe the experiences of women who gave birth in a hospital during the early phases of the COVID-19 pandemic (March–July 2020).

## Methods

We conducted a cross-sectional survey of women 18 years or older who gave birth in a hospital during the COVID-19 pandemic. The primary aim of this study was to describe the experiences of women giving birth during the COVID-19 pandemic. A secondary objective of the study was to identify characteristics correlated with reporting a positive COVID-19 test. Ethical approval was provided by the IRB at the University of *redacted* Medical Center.

Women were recruited via social media (Facebook, Instagram) and word of mouth with study information flyers and ad prompts such as, “What was it like to give birth during a pandemic?” To be included in the study, women had to be between 18 and 50 years old, had to be able to read and write in English or provide their own translator to help them complete the survey, and had to have given birth in a hospital in the United States on or after March 1, 2020. Those who were excluded included men, women who gave birth prior to the onset of the COVID-19 pandemic, and women who gave birth outside of a hospital.

Informed consent and study data from an 80-item survey were collected and managed using REDCap electronic data capture tools hosted at the University of *redacted* Medical Center. REDCap (Research Electronic Data Capture) is a secure, web-based software platform designed to support data capture for research studies [17-18].

Data analysis was completed using Stata 16.1 [19]. Descriptive maternal and birth characteristics were stratified by route of delivery and reported overall. Univariate chi-squared tests were used to test whether there was a correlation between categorical participant or pregnancy characteristics and COVID-19 positivity. Univariate Student t-tests were used to test the possible association between continuous variables and COVID-19 positivity. Multivariable analysis was not performed due to the cross-sectional study design and limitations to determining temporal relationship between predictors and outcome as well as the low frequency of COVID-19 in the study population.

## Results

A total of n=885 women were included in the study. As displayed in Table 1, women were predominantly White Non-Hispanic, married, fully employed, and owned their own home. A slight majority of women (55.7%) were multiparous. Women were 18 to 43 years of age, with an average of 29.8 years old with a standard deviation of 4.9. The majority (70.7%; 626/885) of women had a vaginal delivery with a significant minority (29.3%, 259/885) undergoing cesarean delivery.

The majority of mothers in our sample, 98.9% (n=875/885), reported that they either were not tested or tested negative for COVID-19 during pregnancy or delivery, or postpartum. Only 1.13% of mothers (n=10/885) reported testing positive. All newborns in our sample reportedly did not test positive for COVID-19. During univariate analysis, potential risk factors for COVID-19 positivity among mothers included Asian race/ethnicity as displayed in Table 2.

As displayed in Table 3, other positively correlated birth characteristics (p < 0.05) included cesarean delivery and not having a birth partner present at birth. In addition, it was observed that women with a positive coronavirus test tended (p=0.06) to breastfeed for less than 6 weeks.

## Discussion

Our study of 885 women who gave birth in the United States during the COVID-19 pandemic had several interesting findings. Of note regarding overall health, women in our study were more likely to have high blood pressure (22.5%) compared to previous studies which have shown the rate of hypertension in pregnancy to be under 10% [15]. Women in our sample showed high levels of anxiety (33.8%) compared to previous studies reporting anxiety prevalence in pregnancy around 20% [12]. Women in our study also reported depression at a high rate (18.6%), compared to the 12.7% rate typically accepted, although there are other sources with depression in pregnancy rates consistent with our sample [13-14, 20].These symptoms have potential relationships to stress and were no more prevalent in women who had a diagnosis of COVID-19. One could conjecture that women who gave birth and brought an infant into the world during a pandemic may be vulnerable to physical symptoms of stress, regardless of whether they were personally diagnosed with COVID-19.

While most women reported they still felt safe giving birth in the hospital during a pandemic, 20.5% of the women reported that they felt unsafe in the hospital. While we don’t have an exactly comparable study, in one pre-pandemic survey on the perceived safety of place of birth in pregnant women in their third trimester, 12.6% of women reported they would feel safest giving birth outside of a hospital, even when not planning an out of hospital birth [21]. Adding to this, the majority of our sample (61%) reported that they had inadequate birth support during their delivery. Most women were able to have a birth partner present with them, but many women use multiple sources of support when they are giving birth, which may include doulas, friends, or family members. While hospital birth is considered safe from a healthcare perspective, and professional organizations have emphatically affirmed this, pandemic-related maternity care practices may not align with a woman’s perceptions of safety and control [22]. While following public health directives, healthcare policy makers should consider ways to improve the experiences of birthing women and ways to help them feel safe and supported.

The small number of women who were diagnosed with COVID-19 in our study were more likely to undergo cesarean delivery compared to women not diagnosed with COVID-19. While vaginal delivery is not contraindicated in COVID-19 infection, this outcome may be related to early uncertainties about the safety of vaginal birth, or that these women were more critically ill and too unstable to undergo vaginal delivery [2, 23]. Another possibility is that some of these women contracted COVID-19 while hospitalized for childbirth, and having a cesarean may have been a risk factor for contracting COVID-19.

Women with COVID-19 were less likely to report having a birth partner present with them at delivery compared to women without COVID-19, which is consistent with common “no visitor” policies for COVID-19 patients. Additionally, women with COVID-19 tended to breastfeed for less than six weeks. While numerous factors can contribute to breastfeeding length of time, such as illness, milk supply, breast infection, anemia, etc., we found it interesting that women with COVID-19 were just as likely to initiate breastfeeding as their non-COVID-19 infected counterparts. Despite conflicting recommendations between the WHO and CDC, women in our sample who were COVID-19 positive were not separated from their infants, held their infants skin-to-skin after birth, and roomed in with their infants. These behaviors are consistent with the most up-to-date CDC and WHO recommendations, which have wavered and changed throughout the pandemic [4-6, 24] Infants in the non-COVID-19 groups showed no difference in term delivery. While it was statistically significant that women with COVID-19 were more likely to be of Asian race compared to the general sample, less than 3% of our sample (n=2) was Asian, making this finding possibly random chance.

### Limitations

There were several limitations in this study. First, our sample was recruited through social media and word of mouth, resulting in a sample not representative of the general population. This study collected self-reported data, which is naturally biased, including reports on retrospective experiences, some provided days after giving birth and others several months later. The length of time since giving birth and when a participant completed the study also affected how they reported the length of time they breastfed. The outcomes on women who reported testing positive for COVID-19, even when statistically significant, are unreliable due to the small sample size and not specifically separating COVID-19 positivity into during pregnancy, childbirth, or postpartum.

### Recommendations

Just as the COVID-19 virus is novel, the experiences of giving birth in the US during a pandemic of this magnitude are novel. Further research should be conducted prospectively on all women giving birth during a pandemic, focusing on both biological and psychosocial variables. The elevated levels of anxiety, depression, and hypertension in our sample deserve closer study. Future research should also focus on race, ethnicity, and socioeconomic variables and how those may affect health and psychosocial outcomes in birthing mothers during a pandemic.

Several registries of women positive with COVID-19 have been created and will hopefully begin addressing many health outcome questions in large samples of infected women [25-27]. Since we can’t determine causation in our study, future research should look at the relationships between cesarean delivery, breastfeeding, race, and COVID-19 infection in pregnant women.

Healthcare policy and maternity care practices should focus not only on keeping women safe from COVID-19 infection, but also on ways to increase women’s overall feelings of safety and control in their birthing environment. As professional organizations affirm that the hospital is safe for labor and delivery even during a pandemic [11], the definition of safety should be broadened to include psychological safety. A positive healthcare maternity care experience for a woman giving birth in a hospital, even during an unprecedented pandemic, can have a lasting impact on how she and her family interact with healthcare for the rest of their lives.

### Conclusion

Our study of 885 women who gave birth during the early phase of the COVID-19 pandemic showed that the women reported high levels of anxiety, depression, and hypertension.

The majority of women reported inadequate support in labor, and a relatively high number reported they did not feel safe giving birth in a hospital.

A small portion of our sample reported testing positive for COVID-19 either during pregnancy, during childbirth, or postpartum. Women who tested positive for COVID-19 were more likely to be of Asian race, have a cesarean, and discontinue breastfeeding before six weeks than non-COVID-infected women.

Future research should focus on the biological and psychosocial variables surrounding giving birth during a pandemic. Healthcare policy and maternity care practices should focus on improving the feelings of safety and control and overall experience for women giving birth in the hospital during a pandemic.

## Data Availability

The data that support the findings of this study are available from the corresponding author, upon reasonable request.

## Notes

### Competing Interest Statement

The authors have declared no competing interest.

### Funding Statement

This study was funded by the Sigma Theta Tau International, Gamma Pi Chapter, Sister Patricia Miller Award

### Author Declarations

IRB approval was obtained through the University of Nebraska Medical Center. IRB #303-20-EP

